# Targeted Sequencing of CTX-M Alleles in Seattle Area Wastewater

**DOI:** 10.1101/2024.05.24.24307913

**Authors:** Angelo Q. Ong, Sarah E. Philo, Anysiah Taylor, Ruohan Hu, John Scott Meschke, Erica R. Fuhrmeister

**Author notes:** Correspondence to: Dr. Erica Fuhrmeister, 4225 Roosevelt Way, Seattle, WA 98105.

## Abstract

Extended-spectrum-beta-lactamases (ESBLs) are a growing group of antimicrobial resistance (AMR) enzymes that can result in severe clinical outcomes. The CTX-M gene, which encodes for ESBLs in bacteria, confers resistance to third generation cephalosporins and is of high clinical concern. We developed a targeted, long-read sequencing method utilizing unique molecular identifiers to generate accurate, full length CTX-M gene sequences from wastewater. We characterized CTX-M in 36 samples from three Seattle area wastewater treatment plants from April 2020 to March 2021. We identified a core community of alleles that persisted across time and treatment plant. The CTX-M-15 containing protein variant (CTX-M-15/216/28) was detected in all but three samples and made up, at most, 30% of detected CTX-M alleles. We observed significant diversity across the CTX-M gene at the nucleic acid level, although most nucleotide mutations were synonymous - resulting in two to three amino acid variants across 19 loci. By average relative abundance, 23% of protein variants were novel, defined as those not represented in the CARD database. This method provides information (full length gene sequences) that cannot be obtained through other culture-independent methods. This flexible approach can be expanded to additional targets and implemented in settings where AMR surveillance is a priority, such as hospital wastewater.

## Introduction

The emergence of antimicrobial resistance (AMR) is an existential threat to human and animal health. In 2019, 4.95 million human deaths were associated with bacterial antimicrobial resistance.^1^ The expansion of extended-spectrum beta-lactamases (ESBLs) in multiple genera of bacteria has further worsened the landscape of antibiotic resistance.^2^ ESBLs are enzymes that confer resistance to a wide array of beta-lactam antibiotics targeting the bacterial cell wall. CTX-M (cefotaximase-München-lactamase) is an antibiotic resistance gene (ARG) that encodes for ESBLs and has high efficacy against third generation beta-lactam cephalosporins (3GC). 3GCs are primarily reserved for difficult-to-treat infections such as sexually transmitted infections,^3^ neonatal sepsis, and meningitis.^4^ Resistance to 3GCs such as ceftriaxone has been observed in many taxa including *E. coli* and *Salmonella* bacteria harboring CTX-M genes.^5,6^ CTX-M is a highly diversified gene with different alleles conferring varying levels of resistance to beta-lactams. A notable example is CTX-M-15, a protein variant that has increased activity against the 3GC ceftazidime compared to other CTX-M alleles.^7,8^

CTX-M is globally distributed with predominant alleles differing across geographic regions. CTX-M-15 is the most dominant allele in most regions around the world with exceptions to CTX-M-14 in East Asia and CTX-M-2 in South America.^9^ Although there is limited clinical literature for CTX-M in Africa compared to other continents^9^, CTX-M-15 is the most prevalent allele described.^10^ Clonal relationships have been demonstrated in virulent ST131 *E. coli* isolates containing CTX-M-15 in eight countries across three continents.^11^ Despite its clinical significance, CTX-M is not limited to clinical settings as has been detected in a variety of veterinary,^12^ agricultural,^13,14^ and environmental settings.^15,16^

As new CTX-M alleles, subgroups, and clusters are continuously being identified in bacterial isolates,^17–20^ there is a need to characterize alleles at the population-level. Surveillance of ARGs in populations can be done through wastewater-based epidemiology (WBE). WBE has been used to characterize circulation of pathogens and ARGs at different scales (e.g. city, neighborhood, building).^21,22^ ESBLs have been surveilled in wastewater through culture-based, PCR, and DNA sequencing methods.^23–25^ Metagenomic sequencing is commonly used to provide broad characterization of all ARGs found in wastewater. However, current metagenomic methods are prone to oversampling highly abundant ARGs and can lack sensitivity.^26,27^ In addition, short-read sequencing methods (approx. 300 bps) are unable to provide adequate breadth of coverage for ARGs, such as CTX-M, that are approximately 900 base pairs in length. Several clinically significant, low abundant ARGs, such as CTX-M and their alleles, are often overlooked in wastewater. Targeted sequencing increases sequencing depth of low abundance targets and long-read sequencing allows for high breadth of coverage of ARGs.

A notable limitation of long-read sequencing methods (i.e. Nanopore) is the high error rate. Unique molecular identifiers (UMIs) have previously been developed for full length 16S long-read sequencing. High error rates are corrected by tagging template molecules with UMIs during PCR. Then, UMIs in the sequenced amplicons are clustered and binned to generate consensus sequences.^28^ The resulting consensus sequences are highly accurate, span the length of the gene, and enable characterization at the single nucleotide level. In this study, we first developed a targeted, long-read sequencing method that utilizes UMIs to sequence the clinically relevant ARG, CTX-M in wastewater and then demonstrate the utility of this method by characterizing the diversity of CTX-M alleles in Seattle-area wastewater.

## Methods

### Wastewater collection, concentration, and extraction

Primary influent wastewater was grab sampled weekly from three Seattle-area wastewater treatment plants (WWTPs) between April 2020 to March 2021 as part of a previous study.^29^ All grab samples were transported on ice and stored at 4°C prior to processing. For Brightwater WWTP, multiple grab samples from different time points for a single day were composited and mixed well prior to concentration. For the other WWTPs, one grab sample was collected for a single day. All samples were processed and concentrated within one week of the collection date.

0.5 L of primary influent wastewater was concentrated using 5% skimmed milk, shaken, centrifuged, and pellets resuspended in 6 mL of 1X PBS. DNA and RNA were coextracted from 280 μL of concentrated sample using a QIAmp Viral RNA Mini extraction kit (Qiagen) with no DNase treatment. One sample from the first week of each month for 12 months was analyzed for CTX-M variants across the three WWTPs (36 samples total).

### Gene Quantification

qPCR was performed on wastewater nucleic acid extracts to quantify the gene copies (gc) of CTX-M in each sample (alleles captured in Table S1). CTX-M quantities were used to dilute samples to 5000 gc input in Unique Molecular Identifier PCR (described below). A final concentration of 0.9 μM for forward and reverse primers, 0.25 μM of probe, and 1X of TaqMan Fast Advanced MM (Invitrogen) was used in each reaction (Table S2). 2 μL of sample was used with 18 μL of qPCR mastermix and samples were run on a StepOne Plus (Applied Biosystems). Undiluted and ten-fold diluted samples were run in duplicate. A gBlock (Integrated DNA Technologies) was used to generate a standard curve, from 10 to 10^6^ gc per reaction, in triplicate. Triplicate negative controls with 18 μL of qPCR mastermix and 2 μL of molecular grade water were included. Cycling conditions for all thermocycling are available in Table S3.

### Unique Molecular Identifier PCR

To add unique molecular identifiers (UMIs) to CTX-M genes and amplify the gene and UMIs, two sequential PCR reactions were performed (**Figure 1**). The first reaction used two cycles and primers targeting the CTX-M gene contained an 18 nt random sequence (Table S2). 5000 gcs of CTX-M were used as input to the PCR reaction for a total reaction volume of 25 μL (qPCR results in Figure S1). The mastermix consisted of 0.2 μM forward and reverse CTX-M primer, 0.5 mM dNTPs, 1X SuperFI II buffer (Invitrogen), 1U SuperFI II DNA polymerase, and molecular grade water. 930 bp PCR products were cleaned using 0.9X magnetic beads (Sergi Lab Supplies) with 80% ethanol.

**Figure 1:**
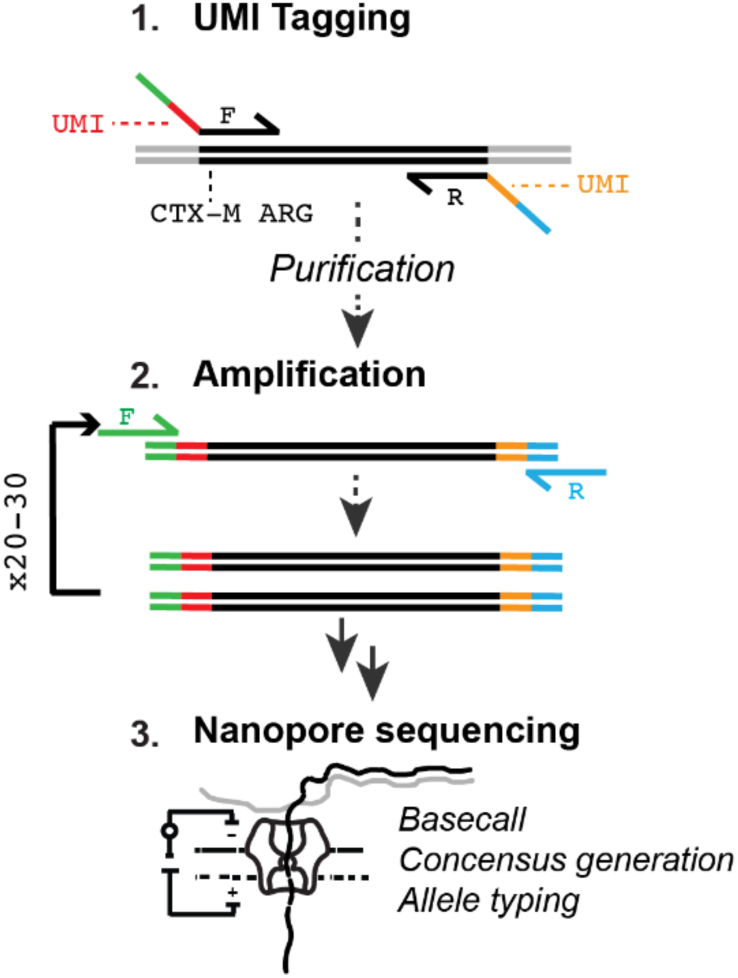
Schematic of UMI PCR Protocol. UMIs are added to each template molecular. Tagged molecules are subsequently amplified and sequenced. Consensus sequences are generated from binning by UMIs.

The second reaction of UMI PCR was performed with primers to amplify the UMIs along with the template molecules. Mastermix consisted of 0.2 μM forward and reverse primers, 0.5 mM dNTPs, 1X SuperFI II buffer, 1U SuperFI II polymerase, and molecular grade water. 20 μL of the previous PCR product was used as input DNA for a total reaction volume of 50 μL. PCR products were loaded with 6X TriTrack DNA Loading Dye (ThermoScientific) and amplification was visualized on 1% agarose gels stained with GelGreen (Biotium). PCR products were cleaned as described above.

PCR products were subsequently barcoded to sequence multiple samples on a MinION (Table S4). The final concentration of mastermix for barcoding PCR consisted of 0.2 μM forward and reverse primer, 0.5 mM dNTPs, 1X SuperFI II buffer, 1U SuperFI DNA polymerase, and molecular grade water. 20 μL of the previous PCR product was used as input DNA for a total reaction volume of 100 μL. All PCR thermocycling was performed on a T100 thermal cycler (Bio-Rad Laboratories). PCR products were cleaned as described above. For two of six batches, a ratio of 0.5X beads was used to exclude low molecular weight DNA bands. DNA concentrations for PCR products were measured using the Qubit 1X dsDNA High Sensitivity assay kit (Invitrogen) on a DeNovix Fluorometer and then 70-100 fmols were pooled for sequencing.

### DNA Sequencing

Pooled PCR products were prepared for sequencing using Oxford Nanopore’s ligation sequencing kit (SQK-LSK-114) according to the manufacturer’s protocol. After adaptor ligation, short fragment buffer and a bead ratio of 0.8X was used in the final cleanup step. R10.4.1 flow cells were run on a MinION MK1B for 36 hours. Sequencing reads are available in NCBI under BioProject PRJNA1107162.

### Data Analysis

Fast5 files were basecalled with guppy (v6.5.7) using the R10.4.1 super accuracy model (dna_r10.4.1_e8.2_400bps_sup.cfg) with a minimum quality score of 9. Basecalled reads were demultiplexed with cutadapt (v2.6)^30^ to search for forward and reverse barcodes at each read end with the following parameters: an error rate of 0.15, a minimum length of 800 bps, and maximum length of 1100 bps. Demultiplexed reads were run through an existing ssUMI pipeline (v0.3.2)^28,31^ to produce consensus sequences based on dual UMIs. The longread_umi ssumi_std command was used with the following parameters: -s 200, -e 200, -E 0.1, -m 800, -M 1100, -f AAGGTTGGCCAGGCTACCCAAAAC, -F CGACGCTAATACATCGCG, -r CAAGCAGAAGACGGCATACGAGAT, -R ATGGTTAAAAAATCACTGCGCCAGT, -c 3, -p 2. In brief, USEARCH (v11.0.667)^32^ was used to identify UMI clusters at the beginning and end of each read. Reads were then grouped by UMI pairs and chimeras were removed. Grouped reads were polished using cycles of Racon (v1.4.10)^33^ and Medaka (v0.11.5)^34^ resulting in a single consensus sequence for each UMI pair bin.

ARGs were identified in resulting consensus sequences using Minimap2 (v2.22)^35^ to map against the Comprehensive Antibiotic Resistance Database (CARD) (v3.2.6).^36^ Nucleotide alleles and amino acid variants were named based on the closest database match in CARD. If sequences mapped at 100% identity, the name of the allele in CARD was retained. If sequences did not map at 100% identity to any CARD sequence, the name of the closest match (highest percent identity) was retained with a “variant” suffix. More than one allele was included in the name if sequences aligned equally to multiple database matches. Alleles that occurred only once in the entire dataset were removed from the analysis to avoid spurious associations. Data were visualized and analyzed in R (4.2.3)^37^ with phyloseq (1.42.0).^38^ Alignment of alleles, frequency plot, and phylogenetic tree were generated using Jalview (2.11.3.2).^39^

To test for differences in alpha diversity in WWTPs, we used a Kruskal-Wallis test to compare the median number of unique alleles and Shannon diversity indices. Flow values, measured in millions of gallons per day (MGD) at each sample date were used to categorize samples as either high or low flow. Samples with a flow value lower than the median were categorized as low flow while samples with a flow value greater than the median were categorized as high flow (West Point median: 67 MGD, Brightwater: 16 MGD, South Plant: 62 MGD). We used a Wilcoxon-Ranked Sum test to determine if there was a statistical difference in alpha diversity between samples collected during high and low flow.

To validate performance of the PCR, sequencing, and data analysis methods, we performed an experiment where two alleles with 1 bp difference were spiked into wastewater and underwent the processing pipeline described above. Our analysis pipeline correctly identified the spiked-in alleles at the single nucleotide level (Figure S2 and additional method details in the supporting information).

## Results and Discussion

### Variant Relative Abundance and Diversity

We observed a core composition of highly abundant CTX-M protein variants across all three WWTPs (**Figure 2A**). The CTX-M-22/3/211 nucleotide allele and CTX-M-22/3 protein variant were the most abundant regardless of time and WWTP (Figure 2A and Figure S3). CTX-M-22 was also detected previously in a study of clinically significant (isolated from a sterile body site or a significant quantity of growth from nonsterile sites) *E. coli* isolates from pediatric patients in Seattle.^40^ The second most abundant protein variant (CTX-M-3/22/30/12 var.) was novel, defined as those that did not map 100% to the CARD database, and one amino acid/5 bps away from CTX-M-22/3. Protein variants CTX-M-30 and CTX-M-206 were also frequently detected across all WWTPs and sampling dates (average relative abundance (RA) of 8.9 and 4.4%, respectively). In a previous study of municipal wastewater in Southern California using cloning and sanger sequencing, the authors also identified a high relative abundance of CTX-M-3 and CTX-M-30 gene alleles,^41^ suggesting that there may be established alleles of CTX-M in human populations or that are established in wastewater sewer systems.

**Figure 2:**
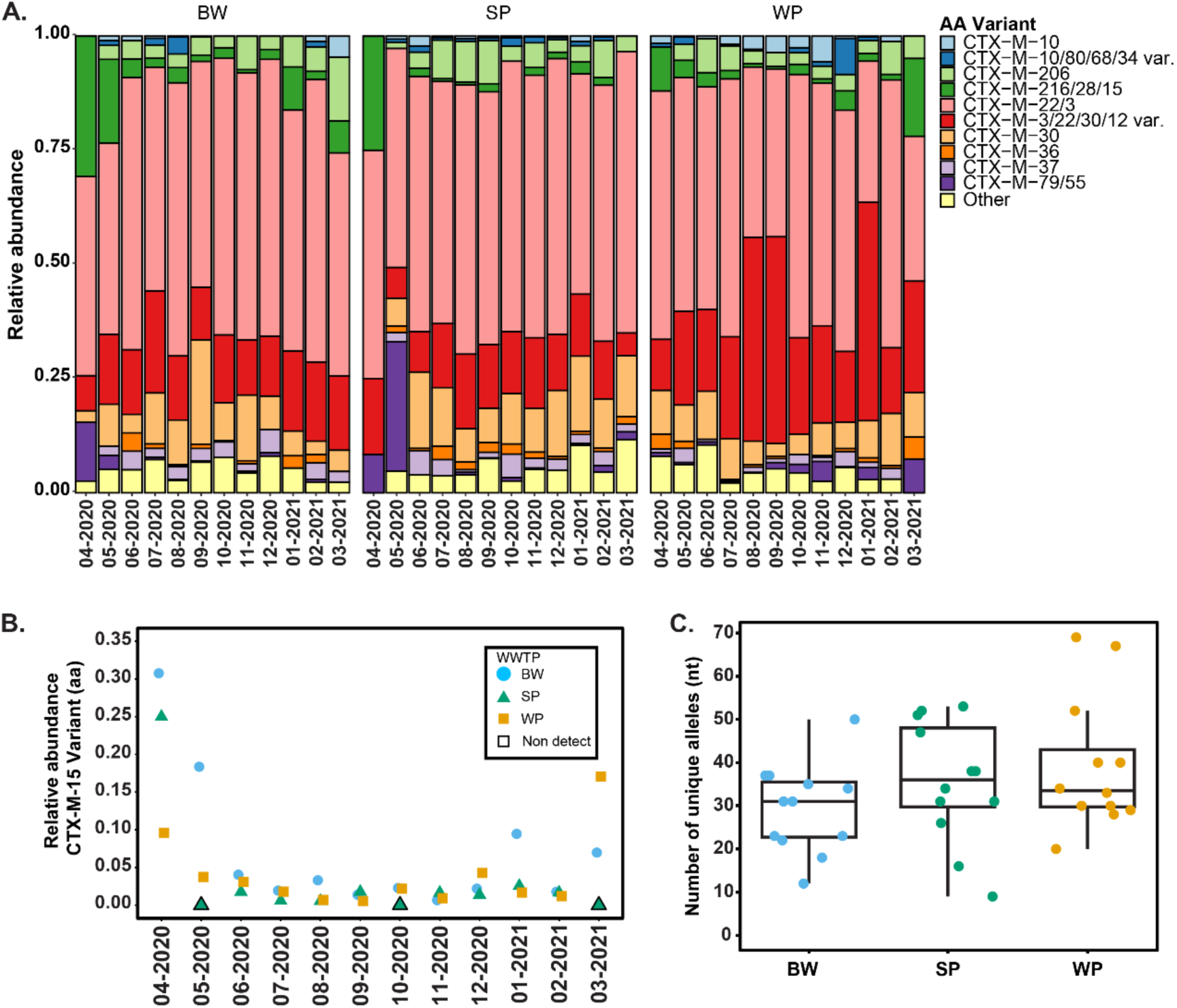
A.) Relative abundance of the top 10 most common protein variants across all samples. B.) Relative abundance of the CTX-M-15 containing protein variant (CTX-M-216/28/15), which has high clinical relevance, across all samples. C.) Number of unique nucleotide alleles by WWTP.

The CTX-M-15 containing protein variant (CTX-M-15/216/28) was detected in all but three samples (SP in May 2020, October 2020, and March 2021). The relative abundance of the CTX-M-15/216/28 protein variant was as high as 30% for BW and 25% for SP in April 2020, measured by consensus sequences. For most samples, the relative abundance was less than 5% of consensus sequences (**Figure 2B**). While CTX-M-15 is globally disseminated, it is also locally significant due to its appearance in both clinical settings and in agriculture. The previously referenced study of clinically significant isolates from Seattle pediatric patients detected CTX-M-15 in 10 out of 49 *E. coli* isolates. The same study also found that most of the CTX-M-15 positive isolates were clones of a ST131 pandemic strain.^40^ In another previous study in Washington state, CTX-M-15 was the most frequently detected allele (50 out of 99 isolates) in CTX-M positive *E. coli* isolates from dairy cattle.^42^

There were no statistically significant differences between WWTPs in the number of unique alleles observed (Kruskal-Wallis p-value=0.40) (**Figure 2C**) or Shannon diversity index (p-value=0.44) (Figure S4) at either the nucleotide or amino acid level (Figure S5). Within each WWTP, there were no significant differences in number of unique alleles/variants or Shannon diversity index between high and low flow (Figures S4 and S5). Each WWTP differs in the population served and land use based on the number of medical facilities, pastureland area, and industrial waste permits (Figure S6 and Table S5). Despite the different contributions to influent wastewater there were no differences in alpha diversity, which may indicate geographically consistent trends.

### Novel Variants in Wastewater

The top 20 CTX-M protein variants, based on relative abundance, were aligned to visualize mutation hotspots (**Figure 3A**). Observed diversity of mutations were confined to 19 discrete loci on the CTX-M gene between residues 13-242 (**Figure 3B**). A subset of these observed mutations is associated with changes in CTX-M function.^43^ For example, mutation D242G in CTX-M-15 has been implicated in conferring increased resistance to ceftazidime.^44^ Residue D242 was previously characterized as a mutation hotspot in CTX-M-1 cluster variants.^18^ We identified two substitutions at this residue: 1.) D242G in CTX-M-79/55 and CTX-M-216/28/15 and 2.) D242N in CTX-M 216/3/22/15 var. Of the mutation hotspots in our work, many have been observed in CTX-M variants from clinical isolates (loci 13, 16, 31,80,112, 212, 242, 45,160),^18^ suggesting our method is capturing true allele diversity.

**Figure 3:**
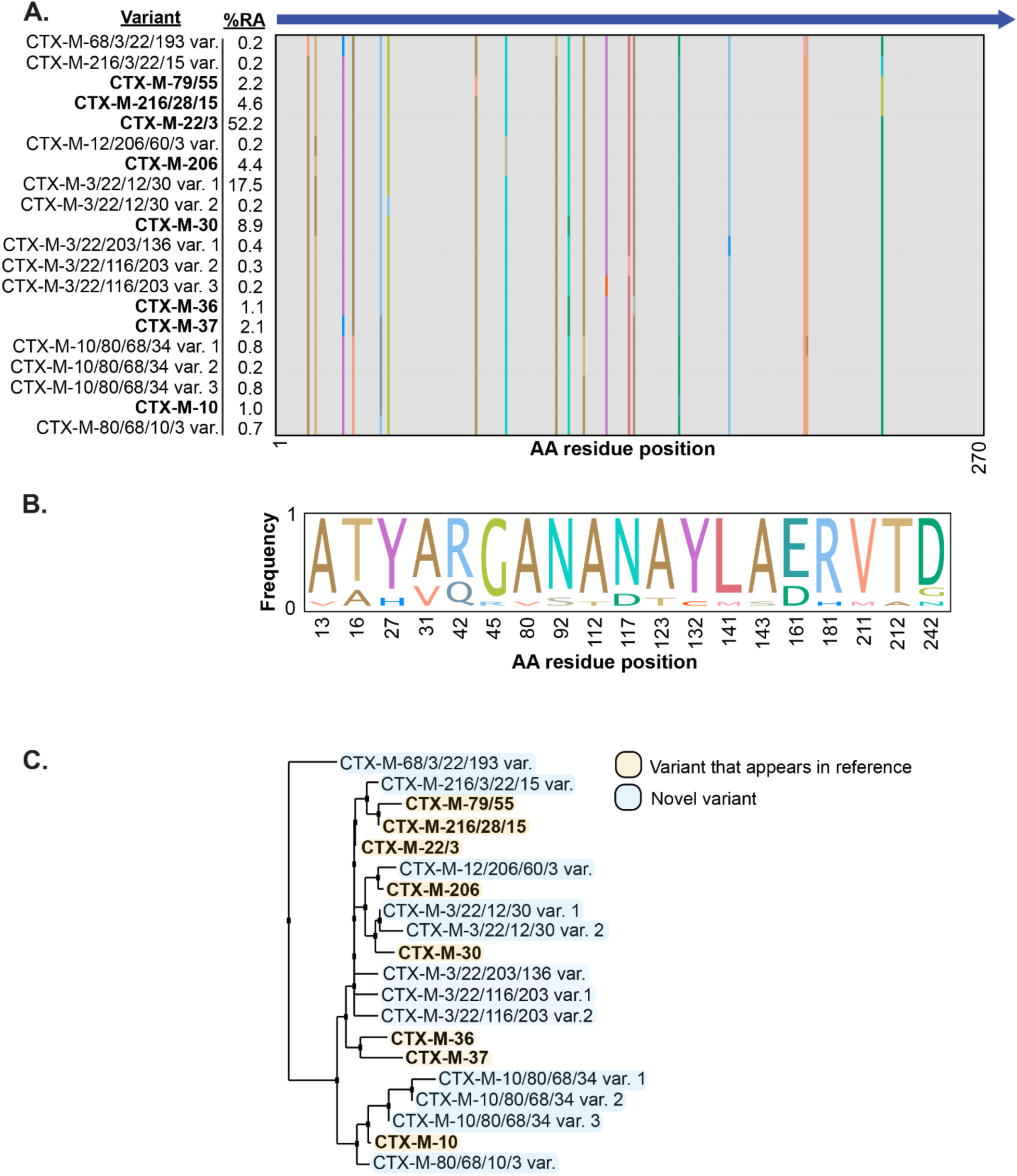
A.) Location of observed mutations across the top 20 protein variants (amino acid level) of the CTX-M gene. B.) Frequency of amino acid substitutions at loci with multiple residues based on unique variants and not abundance weighted C.) Phylogenetic tree of the top 20 protein variants using nearest neighbor joining.

The consensus variant is the most abundant protein variant CTX-M-3/22 (average RA of 52.5%). Despite a larger diversity observed across the CTX-M gene at the nucleic acid level, most nucleotide mutations were synonymous – resulting in just two amino acid variants at all but one position, 242, where we observed three variants. Of these, the majority of amino acid substitutions are neutral (e.g., A13V, T16A) or conservative (e.g., Y27H, R42Q) as classified by BLOSUM62 scoring.^45^ Three were unfavorable or non-conservative (G45R, Y132C, D242G). 12 of the top 20 occurring variants were found to be novel – yet closely match to CTX-M variants in CARD (**Figure 3C**). While many of our protein variants are not observed in CARD, they are observed in other studies of non-clinical isolates. For example, the second most abundant protein variant (CTX-M-3/22/30/12 var. 1) was also found in wastewater.^41^ Similarly, CTX-M-10/80/68/34 var. 1 was found in a *Kluyvera intermedia* isolated from an aquaculture study.^46^ Our work highlights the challenges with using common ARG databases, populated with clinical isolates, when there is an abundance of diverse alleles found in nature that are not represented. Our method offers a rapid approach to surveil the diverse alleles in environmental samples.

### Method Versatility

We demonstrate the utility of this method to characterize CTX-M in wastewater at a resolution that cannot be captured by short-read sequencing. In addition, UMI-based PCR does not necessitate bacterial cloning or culturing to characterize alleles. By coupling allele sequence information with longitudinal data, we can track how CTX-M alleles, and other ARGs-alike, change over time at the population level. Our method is versatile as it can be designed to capture allele clusters or a specific allele depending on the primer design. There is opportunity to multiplex a suite of ARGs for simultaneous allele characterization, including other globally important ESBL targets such as TEM and SHV.^47^

Longitudinal tracking of alleles allows for the establishment of a baseline which can be further leveraged to identify emerging alleles at the population level. Portable allele and variant surveillance can be adapted for other settings such as hospital wastewater and low- and middle-income settings (LMICs). Surveilling hospital wastewater could aid in understanding the diversity of AMR and potentially inform targeted, clinical interventions.^23,48^ In areas with untreated wastewater, such as LMICs, understanding the discharged wastewater ARG alleles can help with public health interventions to reduce proliferation or exposure in the environment.^49,50^ In addition, by characterizing mutation hotspots within ARGs, we can potentially estimate mutations that could result in different phenotypic resistance.

Our study has some limitations. The UMI PCR primers used in this study primarily capture the CTX-M-1 cluster and were not able to identify all CTX-M variants including clinically relevant alleles that have been previously identified locally (e.g. CTX-M-27).^40^ In addition, some alleles were grouped (e.g., CTX-M-15) because they cannot be distinguished from others (e.g., CTX-M-28 or CTX-M-216) due to single base pair mutations outside the reverse primer binding region. Primer design could be optimized with degenerate bases to detect a broader suite of alleles. Finally, we cannot distinguish between anthropogenic or environmental sources of variants. For example, biofilms and bacterial growth in sewage collection systems could contribute to observed variants. There is a critical need to study the relative contributions from environmental versus anthropogenic sources to AMR in wastewater influent.

## Data Availability

Sequencing reads are available in NCBI under BioProject PRJNA1107162.

## Acknowledgements

We thank the local wastewater treatment plants for collecting samples and the UW Royalty Research Fund for support.

## Notes

### Competing Interest Statement

The authors have declared no competing interest.

### Funding Statement

UW Royalty Research Foundation

