## Supplemental Info for "Targeted Sequencing of CTX-M Alleles in Seattle Area Wastewater"

*Correspondence to:

**Table of Contents** **Page**


| Table of Contents | Page |
|---|---|
| Table S1: CTX-M alleles captured by qPCR and UMI primers | 3 |
| Table S2: qPCR and UMI PCR primer sequences | 3 |
| Table S3: qPCR and PCR thermocycling conditions | 4 |
| Figure S1: qPCR gene copies in all samples | 4 |
| Table S4: Barcode sequences | 5 |
| Method Validation | 5-6 |
| Figure S2: Nucleotide-level alleles found in the unspiked wastewater, spiked wastewater, and both samples. | 6 |
| Figure S3: Relative abundance of the top 10 most common alleles across all samples at the nucleotide level. | 6 |
| Figure S4: Alpha diversity at the nucleotide level | 7 |
| Figure S5: Alpha diversity at the amino acid level | 7 |
| Land Use | 8 |
| Figure S6: GIS analysis of WWTPs and their service areas. | 9 |
| Table S5: Medical facility and industrial waste permit counts and pastureland area in the three WWTP service areas. | 9 |

Table S1: CTX-M alleles captured by qPCR and UMI primers.

| Assay | Alleles |
|---|---|
| qPCR primers | CTX-M-155, CTX-M-42, CTX-M-58, CTX-M-88, CTX-M-107, CTX-M-11, CTX-M-101, CTX-M-36, CTX-M-1, CTX-M-30, CTX-M-69, CTX-M-82, CTX-M-117, CTX-M-34, CTX-M-123, CTX-M-55, CTX-M-158, CTX-M-68, CTX-M-108, CTX-M-142, CTX-M-139, CTX-M-61, CTX-M-54, CTX-M-22, CTX-M-114, CTX-M-64, CTX-M-28, CTX-M-33, CTX-M-96, CTX-M-157, CTX-M-109, CTX-M-80, CTX-M-3, CTX-M-52, CTX-M-132, CTX-M-60, CTX-M-103, CTX-M-32, CTX-M-156, CTX-M-71, CTX-M-53, CTX-M-79, CTX-M-66, CTX-M-23, CTX-M-144, CTX-M-12, CTX-M-72, CTX-M-116, CTX-M-136, CTX-M-29, CTX-M-10, CTX-M-37, CTX-M-62, CTX-M-15, CTX-M-162, CTX-M-163, CTX-M-164, CTX-M-166, CTX-M-127, CTX-M-138, CTX-M-143, CTX-M-146, CTX-M-150, CTX-M-153, CTX-M-154, CTX-M-167, CTX-M-169, CTX-M-170, CTX-M-172, CTX-M-173, CTX-M-175, CTX-M-176, CTX-M-177, CTX-M-178, CTX-M-179, CTX-M-180, CTX-M-181, CTX-M-182, CTX-M-183, CTX-M-184, CTX-M-187, CTX-M-188, CTX-M-189, CTX-M-190, CTX-M-193, CTX-M-194, CTX-M-197, CTX-M-199, CTX-M-202, CTX-M-203, CTX-M-204, CTX-M-206, CTX-M-207, CTX-M-208, CTX-M-209, CTX-M-210, CTX-M-211, CTX-M-212, CTX-M-216, CTX-M-218, CTX-M-220, CTX-M-222, CTX-M-224, CTX-M-225, CTX-M-226, CTX-M-227, CTX-M-228, CTX-M-230, CTX-M-231, CTX-M-232, CTX-M-234, CTX-M-236, CTX-M-237, CTX-M-238, CTX-M-244 |
| UMI Primers | CTX-M-101, CTX-M-69, CTX-M-82, CTX-M-117, CTX-M-34, CTX-M-123, CTX-M-55, CTX-M-68, CTX-M-142, CTX-M-139, CTX-M-54, CTX-M-22, CTX-M-114, CTX-M-64, CTX-M-33, CTX-M-157, CTX-M-80, CTX-M-3, CTX-M-52, CTX-M-132, CTX-M-103, CTX-M-156, CTX-M-71, CTX-M-53, CTX-M-79, CTX-M-66, CTX-M-144, CTX-M-72, CTX-M-136, CTX-M-29, CTX-M-10, CTX-M-37, CTX-M-15, CTX-M-127, CTX-M-143, CTX-M-150, CTX-M-153, CTX-M-154, CTX-M-167, CTX-M-170, CTX-M-172, CTX-M-173, CTX-M-176, CTX-M-177, CTX-M-178, CTX-M-179, CTX-M-180, CTX-M-181, CTX-M-182, CTX-M-183, CTX-M-184, CTX-M-186, CTX-M-188, CTX-M-189, CTX-M-190, CTX-M-193, CTX-M-197, CTX-M-199, CTX-M-202, CTX-M-203, CTX-M-204, CTX-M-206, CTX-M-207, CTX-M-208, CTX-M-209, CTX-M-210, CTX-M-212, CTX-M-216, CTX-M-218, CTX-M-220, CTX-M-225, CTX-M-226, CTX-M-227, CTX-M-228, CTX-M-230, CTX-M-231, CTX-M-232, CTX-M-234, CTX-M-236, CTX-M-237, CTX-M-238, CTX-M-244 |

Table S2: qPCR and UMI PCR primer sequences

| Assay | Forward | Reverse | Probe |
|---|---|---|---|
| CTX-M-1 qPCR[1] | 5’-CCGTCACGCTGTTRTTAGGA -3’ | 5’-AATGCCACMCCCAGYCKKCC -3’ | 5’-FAM-CAGCAAAAACTTGCCGRATT -MGB-3’ |
| CTX-M UMI PCR 1 | 5’-CAAGCAGAAGACGGCATACGAGATNNNYRNNNYRNNNYRNNNATGGTTAAAAAATCACTGCGCCAGT-3’ | 5’-AAGGTTGGCCAGGCTACCCAAAACNNNYRNNNYRNNNYRNNNCGACGCTAATACATCGCG-3’ | - |
| CTX-M UMI PCR 2 | 5’-CAAGCAGAAGACGGCATACGAGAT-3’ | 5’-AAGGTTGGCCAGGCTACCCAAAAC -3’ | - |

Table S3: qPCR and PCR thermocycling conditions.

| | | | |
|---|---|---|---|
| qPCR | | | |
| Step | Temperature (°C) | Time | Number of cycles |
| UNG activation | 50 | 2 minutes | |
| Polymerase activation | 95 | 20 seconds | 40X |
| Denaturation | 95 | 15 seconds | |
| Annealing/extension | 60 | 1 minute | |
| UMI PCR 1 (Tagging PCR) | | | |
| Step | Temperature (°C) | Time | Number of cycles |
| Denaturation | 98 | 5 minutes | |
| Denaturation | 98 | 30 seconds | 2x |
| Annealing | 65 | 30 seconds | |
| Extension | 72 | 30 seconds | |
| UMI PCR 2 (Amplification) | | | |
| Step | Temperature (°C) | Time | Number of cycles |
| Denaturation | 98 | 5 minutes | |
| Denaturation | 98 | 15 seconds | 22X |
| Annealing | 65 | 30 seconds | |
| Extension | 72 | 30 seconds | |
| Extension | 72 | 5 minutes | |
| Barcoding | | | |
| Step | Temperature (°C) | Time | Number of cycles |
| Denaturation | 98 | 5 minutes | |
| Denaturation | 98 | 30 seconds | 6X |
| Annealing | 65 | 30 seconds | |
| Extension | 72 | 30 seconds | |

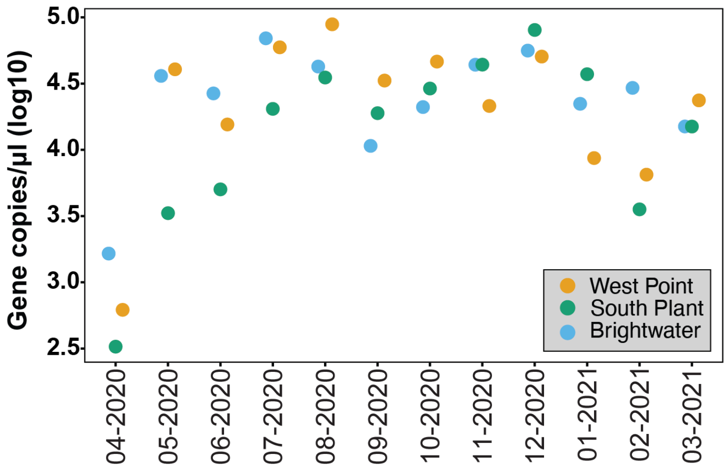


Figure S1: Abundance of CTX-M genes in three Seattle area WWTPs from April 2020-March 2021. qPCR was used to standardize 5000 gene copies of input into unique molecular identifier PCR.

Table S4: Barcode sequences.

| Barcode ID | Primer direction | Sequence |
|---|---|---|
| BC1 | Forward | 5’-CACAAAGACACCGACAACTTTCTTCAAGCAGAAGACGGCATACGAGA -3’ |
| BC2 | Forward | 5’-ACAGACGACTACAAACGGAATCGACAAGCAGAAGACGGCATACGAGA-3’ |
| BC3 | Forward | 5’-CCTGGTAACTGGGACACAAGACTCCAAGCAGAAGACGGCATACGAGA-3’ |
| BC4 | Forward | 5’-TAGGGAAACACGATAGAATCCGAACAAGCAGAAGACGGCATACGAGA-3’ |
| BC5 | Forward | 5’-AAGGTTACACAAACCCTGGACAAGCAAGCAGAAGACGGCATACGAGA-3’ |
| BC6 | Forward | 5’-GACTACTTTCTGCCTTTGCGAGAACAAGCAGAAGACGGCATACGAGA-3’ |
| BC7 | Reverse | 5’-AAGGATTCATTCCCACGGTAACACAAGGTTGGCCAGGCTACCCAAAAC-3’ |
| BC8 | Reverse | 5’-ACGTAACTTGGTTTGTTCCCTGAAAAGGTTGGCCAGGCTACCCAAAAC-3’ |
| BC9 | Reverse | 5’-AACCAAGACTCGCTGTGCCTAGTTAAGGTTGGCCAGGCTACCCAAAAC-3’ |
| BC10 | Reverse | 5’-GAGAGGACAAAGGTTTCAACGCTTAAGGTTGGCCAGGCTACCCAAAAC-3’ |
| BC11 | Reverse | 5’-TCCATTCCCTCCGATAGATGAAACAAGGTTGGCCAGGCTACCCAAAAC-3’ |
| BC12 | Reverse | 5’-TCCGATTCTGCTTCTTTCTACCTGAAGGTTGGCCAGGCTACCCAAAAC-3’ |

**Method Validation**

Wastewater DNA extract was seeded with one *E. coli* isolate and one *K. pneumoniae* isolate that carry different CTX-M alleles with 1bp difference to validate the targeted sequencing workflow. *K. pneumoniae* isolate 0145, containing CTX-M-15, came from the CDC/FDA AR Isolate Bank Enterobacterales Carbapenemase Diversity Panel and the *E. coli,* containing CTX-M-55, was isolated from the stool of a child participating in a 2016-2019 cohort study of enteric infections in Lima, Peru (NIH R01AI108695-01A1). Both isolates were grown for 16 hours in LB broth (BD Difco) in a shaking incubator at 37°C. 5 mL of broth was pelleted by centrifugation at 5000 x g for 5 minutes at 20°C. DNA was extracted from the pellets using a Monarch® Genomic DNA Purification Kit (New England Biolabs). CTX-M was quantified for each DNA extract using qPCR (same protocol as main text). Equal amounts of DNA extracts containing CTX-M-15 and

CTX-M-55 were seeded into wastewater DNA extract and diluted to 5000 gene copies for UMI PCR. The unseeded wastewater sample was also diluted to 5000 gene copies. PCR, sequencing, and analysis were performed as described in the main text.

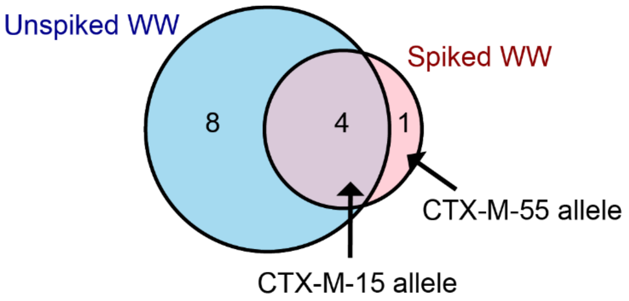


Figure S2: Nucleotide-level alleles found in the unspiked wastewater, spiked wastewater, and both samples.

Eight unique alleles were found in the unspiked wastewater sample only and one unique allele in the spiked wastewater only. Four alleles were common to both samples. We were able to differentiate the spiked alleles (1 bp different) with the CTX-M-15 allele common to both samples and CTX-M-55 present only in the spiked sample.

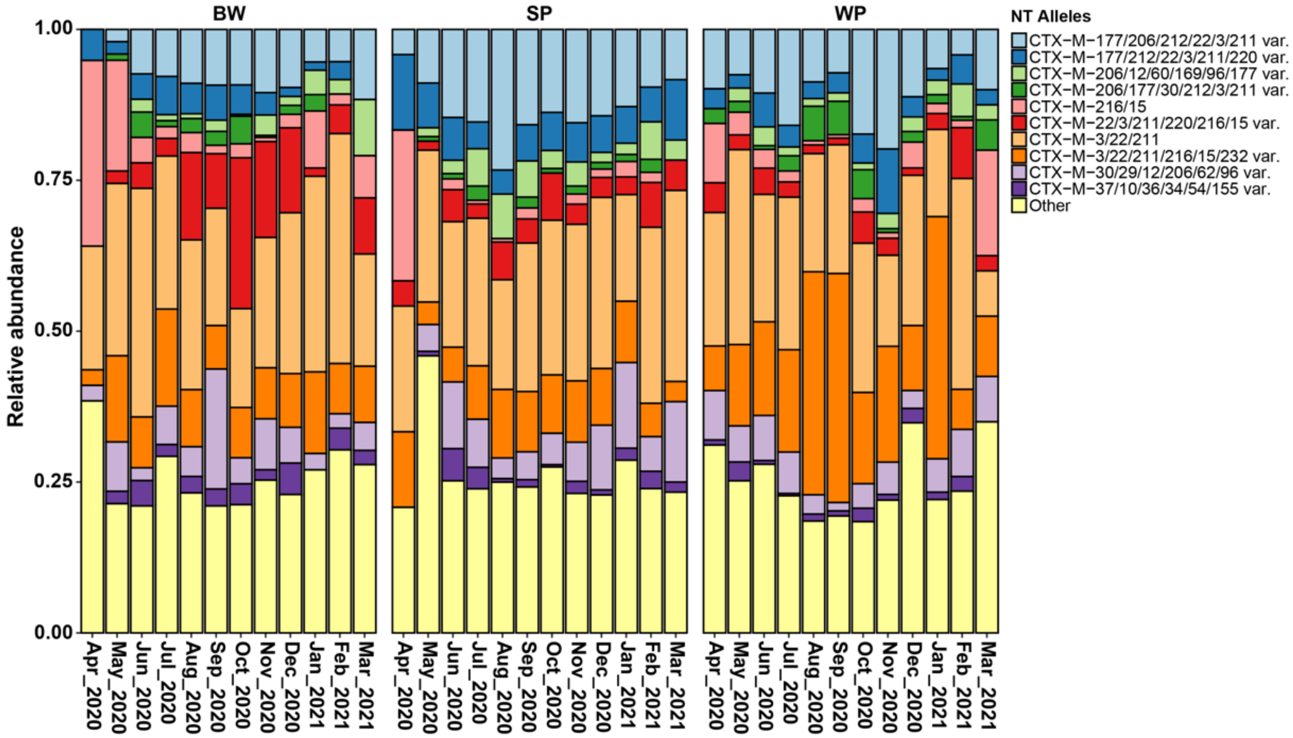


Figure S3: Relative abundance of the top 10 most common alleles across all samples at the nucleotide level.

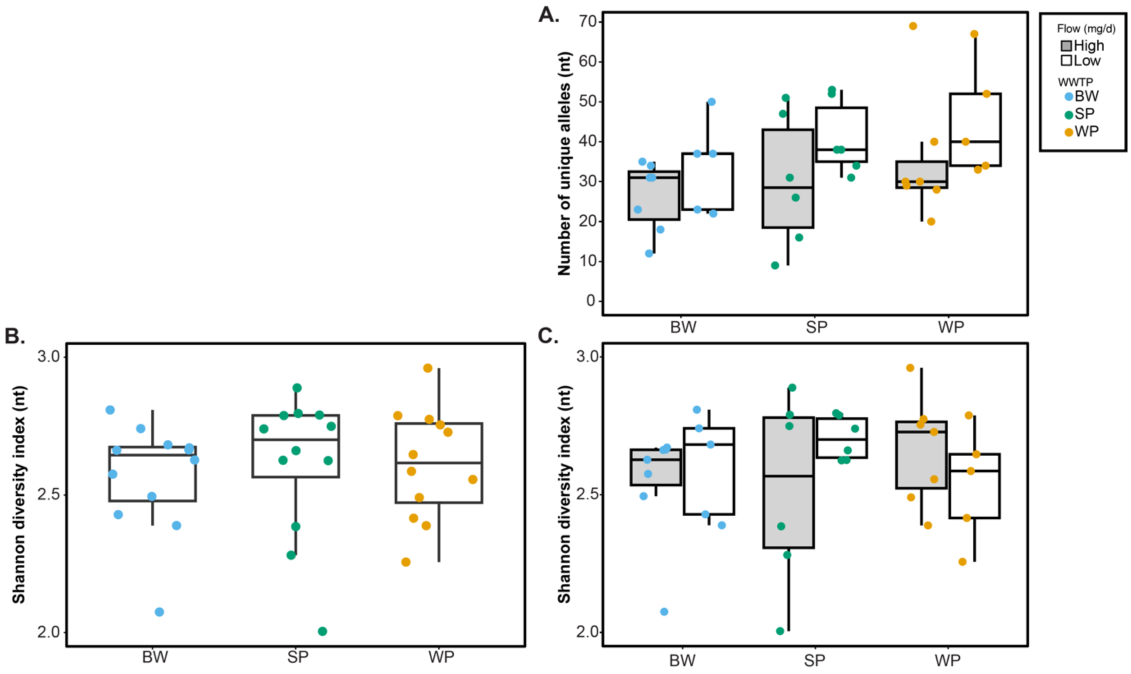


Figure S4: A.) Number of unique alleles by WWTP at the **nucleotide level**, separated by low and high flow. High flow was classified as above the median of the dataset and low flow as below the median of the dataset (per treatment plant). B.) Shannon diversity at the nucleotide level by treatment plant. C.) Shannon diversity, separated by low and high flow.

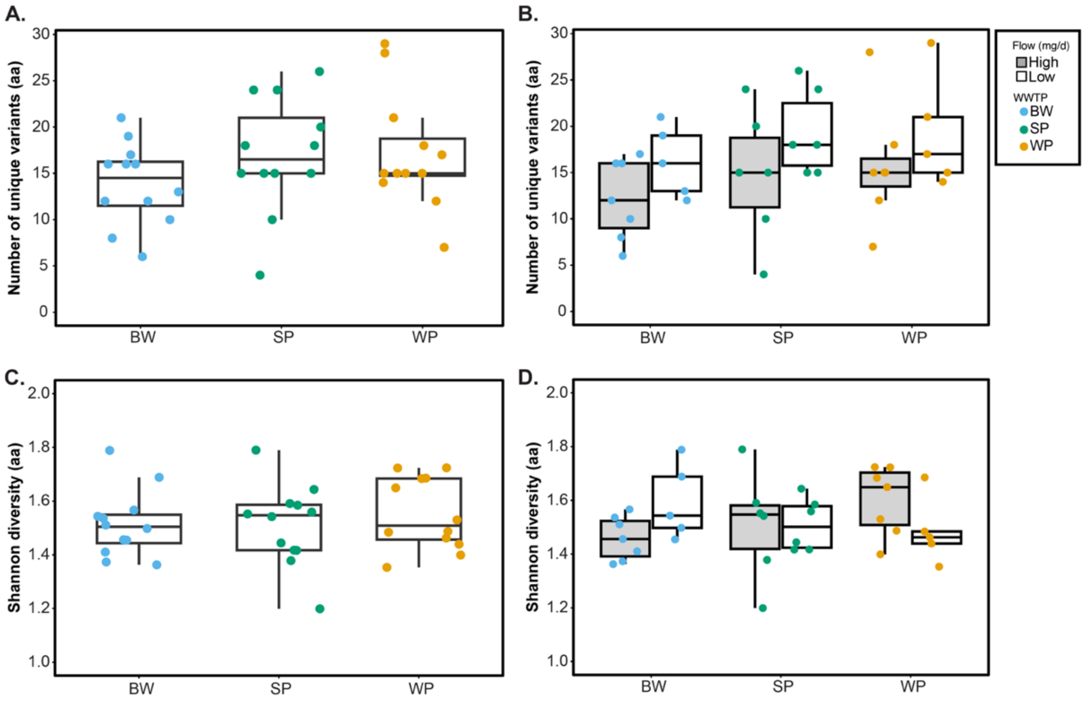


Figure S5: A.) Number of unique alleles by WWTP at the **amino acid** level. B.) Number of unique alleles by WWTP at the amino acid level, separated by low and high flow. High flow was classified as above the median of the dataset and low flow as below the median of the dataset (per treatment plant). C.) Shannon diversity at the amino acid level by treatment plant. D.) Shannon diversity, separated by low and high flow.

**Land Use**

Medical facility points were extracted from King County GIS Open Data and includes facilities such as medical centers, hospitals, and dental centers[2]. Pastureland polygons from the Agricultural Land Use crop data were extracted from the Washington State Geospatial Open Data[3]. Within the crop data, only pastureland was used as a subset since antibiotics are more widely used for livestock animals than for other types of agricultural crops. The industrial waste indicator also came from King County GIS Open Data and includes all industrial facilities that have an industrial waste permit[4]. Industrial waste permits are required for businesses or entities that discharge industrial waste into publicly owned WWTPs[5]. All relevant shapefiles were downloaded and loaded into ArcMap for analysis. New layers were created to select for WWTP service areas and only pastureland from the land use data. Once all layers were loaded, spatial joins were used to quantify the number of medical facilities, and industrial waste points in each WWTP service area. For pastureland, a spatial intersection was performed against the WWTP service area layers and the overlapping area was summed into hectares. Results are summarized in Table S5.

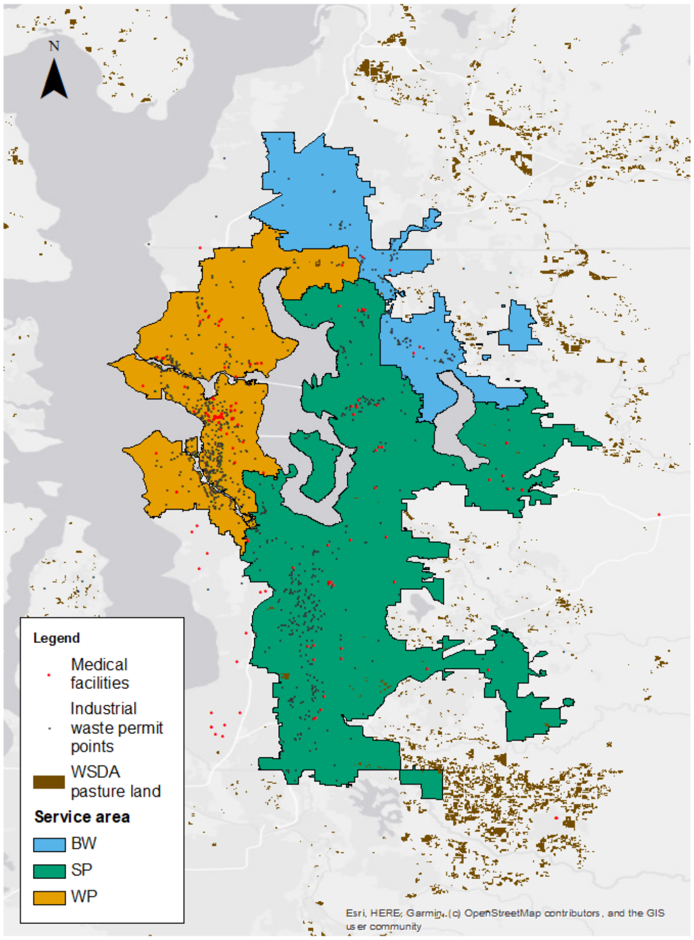


Figure S6: GIS analysis of WWTPs and their service areas.

Table S5: Medical facility and industrial waste permit counts and pastureland area in the three WWTP service areas.

| WWTP | Medical facilities count | Pastureland (hectares) | Industrial waste permit point count |
|---|---|---|---|
| BW | 4 | 16 | 121 |
| SP | 44 | 383 | 376 |
| WP | 60 | 59 | 595 |
